# COVID-19 outbreak in Algeria: A mathematical Model to predict cumulative cases

**DOI:** 10.1101/2020.03.20.20039891

**Authors:** Mohamed Hamidouche

## Abstract

**Introduction:** Since December 29, 2019 a pandemic of new novel coronavirus-infected pneumonia named COVID-19 has started from Wuhan, China, has led to 254 996 confirmed cases until midday March 20, 2020. Sporadic cases have been imported worldwide, in Algeria, the first case reported on February 25, 2020 was imported from Italy, and then the epidemic has spread to other parts of the country very quickly with 139 confirmed cases until March 21, 2020.

**Methods:** It is crucial to estimate the cases number growth in the early stages of the outbreak, to this end, we have implemented the Alg-COVID-19 Model which allows to predict the incidence and the reproduction number R0 in the coming months in order to help decision makers.

The Alg-COVIS-19 Model initial equation 1, estimates the cumulative cases at t prediction time using two parameters: the reproduction number R0 and the serial interval SI.

**Results:** We found R0=2.55 based on actual incidence at the first 25 days, using the serial interval SI= 4,4 and the prediction time t=26. The herd immunity HI estimated is HI=61%. Also, The Covid-19 incidence predicted with the Alg-COVID-19 Model fits closely the actual incidence during the first 26 days of the epidemic in Algeria Fig. 1.A. which allows us to use it.

According to Alg-COVID-19 Model, the number of cases will exceed 5000 on the 42^th^ day (April 7^th^) and it will double to 10000 on 46th day of the epidemic (April 11^th^), thus, exponential phase will begin (Table 1; Fig.1.B) and increases continuously until reaching à herd immunity of 61% unless serious preventive measures are considered.

**Discussion:** This model is valid only when the majority of the population is vulnerable to COVID-19 infection, however, it can be updated to fit the new parameters values.

## Introduction

On December 29, 2019, Wuhan, the capital city of Hubei Province in Central China, has reported four cases of pneumonia with unknown etiology (unknown cause), the next day, the WHO China Country Office was informed (WHO.a 2020) about this pneumonia cases that were found to have a link with Huanan seafood and animal market in Wuhan, the Centers for Disease Control and Prevention (CDC) and Chinese health authorities determined and announced later that a novel coronavirus denoted as Wuhan (CoV) had caused the pneumonia outbreak (CDC 2020). Since then, the outbreak has rapidly spread over a short span of time and has received considerable global attention.

On January 7, 2020 the etiological agent of the outbreak was identified as a novel coronavirus (2019-nCoV) and its gene sequence was quickly submitted (GenBank 2019), the coronavirus was renamed COVID-19 by WHO on February 12, 2020. It has since been identified as a zoonotic coronavirus, similar to SARS and MERS coronaviruses. (Ying, et al. 2020)

On January 30, WHO announced the listing of this novel coronavirus-infected pneumonia (NCP) as a “public health emergency of international concern”, A total of 254 996 confirmed cases of infection with COVID-19, including 10444 deaths have been reported worldwide as on midday March 20^th^, 2020 (WHO.a 2020).

Sporadic cases have been imported to Europe, Africa and North and South America via returning travellers from China. In Algeria, the first case of COVID-19 was reported on February 25, 2020, when an Italian national tested positive in Ouargla region in the south of the country, a few days later, on March 1, 2020, two cases were reported in Blida region in the North of Algeria, following their contacts with two Algerian nationals who came from France for holidays, they were detected positive after their return to France, since then, a COVID-19 outbreak has started in this region (Blida) that form a cluster of more than 5,4 million inhabitants with the surrounding cities (Algiers, Boumerdes, Tipaza) (Algerian Ministry of Health 2020, ONSA 2020), now, the epidemic is spreading to other parts of the country, until March 22, 2020, the Algerian authorities have declared 200 confirmed cases with a fatality rate of 8,5% (17 deaths) (Algerian Ministry of Health 2020).

Meanwhile, there is considerable uncertainty as to the extent of the epidemic and its parameters, the COVOD-19 reproduction number (R0) has been estimated in various studies, they found, 2,35 (95% CI 1.15–4.77) (Kucharski, et al. 2020), two studies used stochastic methods to estimate R0 have reported a range of 2.2 to 2.68 with an average of 2.44. (Joseph, et al. 2020, Riou et Althaus 2020), Six studies that used mathematical methods to estimate R0 produced a range from 1.5 to 6.49 with an average of 4.2. (Shen, et al. 2020, Read, et al. 2020, Chong, et al. 2020, Natsuko, et al. 2020, Tang, et al. 2020) The three studies using statistical methods such as exponential growth estimated an R0 ranging from 2.2 to 3.58, with an average of 2.67. (Liu, et al. 2020, Zhao, et al. 2020). Also, the COVID-19 fatality rate vary by region from 0.39% in Norway to 8,3% in Italy, in China the fatality rate is 4%, (Wilson, et al. 2020, Johns Hopkins 2020), however, the highest mortality rate remain in Algeria at 8,5% until March 22, 2020 (Algerian Ministry of Health 2020)

In the early stages of a new infectious disease outbreak, as COVID-19 in Algeria, it is crucial to estimate the transmission dynamics and inform predictions about potential future epidemic growth (Viboud, et al. 2018), it can provide insights into the epidemiological situation which help decision makers to adapt the health system capacities, thus, a prediction Model can help to do that and identify whether outbreak control measures are having a measurable effect or not (Funk, et al. 2017, Riley, et al. 2003) and guide the design of alternative interventions (Kucharski, et al. 2015), in addition, a prediction Model can be updated to help estimate risk to other countries (Cooper, et al. 2006).

To this end, we have implemented the Alg-COVID-19 Model which allows to predict the cumulative cases in the coming weeks, and to calculate the actual basic reproduction number (R0), consequently, this Model can show hospitals what to expect in terms of Covid-19 patients, the percentage who need to be in an intensive care unit (ICU) or on a ventilator and the future number of deaths based on a given data.

## Methods

### Epidemiological data

We retrieved information on cases number with confirmed COVID-19 infection based on official reports from governmental institutes in Algeria (Algerian Ministry of Health 2020).

### The mathematical Model

In order to predict the cumulative cases of COVID-19 Algerian epidemic in the coming weeks we used the mathematical model (Alg-COVID-19) defined as:

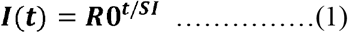

**I(t):** The incidence (cases number) at t time

**R0:** The reproduction number

**SI:** The serial interval

**t:** The prediction time

### Estimation of Basic Reproduction Number (R0) and the herd immunity

The basic reproduction number is defined as the (average) number of new infections generated by one infected individual during the entire infectious period in a fully susceptible population. It can be also understood as the average number of infections caused by a typical individual during the early stage of an outbreak when nearly all individuals in the population are susceptible to infection. The basic reproduction number reflects the ability of an infection spreading under no control, it has three components (Roy, et al. 1992):

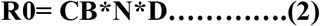

CB: effective contact rate (C: contact frequency, B: contact efficacy)

N: susceptible population size

D: infectious phase duration

The approach implemented to estimate the basic reproduction numbers (R0) in this model is to calculate the average R0 using the actual chronological cumulative cases during the first 26 days of the COVID-19 Algeria epidemic between February 25 and March 22, 2020, so that, we used the equation (3) derived from the equation (1). The R0 will be used also to calculate the herd immunity (HI) needed to stop the epidemic spontaneously based on the equation 4.

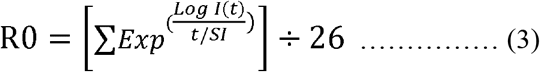

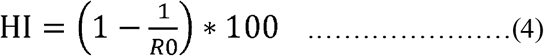

### Estimation of the serial interval (SI)

The serial interval (SI) is the time between symptom onset of a primary and secondary case, a previous studies reported that (SI = 4.4±3.17) days (Chong, et al. 2020), so we used this value in our model to predict the incidence.

### Statistical analysis and software

This study was conducted using Excel 2013 and STATA/IC 15 software

## Results

### The initial parameters used in Alg-COVID-19 Model

Without considering the prevention measures and other factors, this paper focused on the predicted COVID-19 incidence at any time of the epidemic using Alg-COVID-19 Model (equation 1), based on three parameter combinations that create plausible epidemic curves, namely, the average basic reproduction number calculated with the actual cases number at the first 26 days R0=2.55 (95% CI 2.15–2.93) (equation 3), the serial interval (SI=4.4) and the prediction time (t) we are looking for. The herd immunity (HI) estimated using the (equation 4) is HI=61%.

### Model fitting

The Covid-19 incidence (cases number) predicted with the Alg-COVID-19 Model fits closely the actual incidence during the first 26 days of the epidemic in Algeria (Fig. 1.A), this allows us to use this Model to predict the Covid-19 incidence in the next months of the epidemic (Fig. 2.B).

**Table 1.** Alg-COVID-19 Model results for next weeks

| Date | Days | Cases | Date | Days | Cases | Date | Days | Cases |
| --- | --- | --- | --- | --- | --- | --- | --- | --- |
| 21/03/2020 | 25 | 139 | 12/04/2020 | 47 | 14881,5074 | 04/05/2020 | 69 | 1593232,1 |
| 22/03/2020 | 26 | 171,897993 | 13/04/2020 | 48 | 18403,6061 | 05/05/2020 | 70 | 1970312,24 |
| 23/03/2020 | 27 | 212,582158 | 14/04/2020 | 49 | 22759,3019 | 06/05/2020 | 71 | 2436638,26 |
| 24/03/2020 | 28 | 262,895297 | 15/04/2020 | 50 | 28145,8871 | 07/05/2020 | 72 | 3013332,57 |
| 25/03/2020 | 29 | 325,116359 | 16/04/2020 | 51 | 34807,3489 | 08/05/2020 | 73 | 3726516,7 |
| 26/03/2020 | 30 | 402,063666 | 17/04/2020 | 52 | 43045,4203 | 09/05/2020 | 74 | 4608494,54 |
| 27/03/2020 | 31 | 497,22257 | 18/04/2020 | 53 | 53233,2472 | 10/05/2020 | 75 | 5699215,55 |
| 28/03/2020 | 32 | 614,903323 | 19/04/2020 | 54 | 65832,2902 | 11/05/2020 | 76 | 7048084,27 |
| 29/03/2020 | 33 | 760,436309 | 20/04/2020 | 55 | 81413,2271 | 12/05/2020 | 77 | 8716198,13 |
| 30/03/2020 | 34 | 940,413492 | 21/04/2020 | 56 | 100681,801 | 13/05/2020 | 78 | 10779114,9 |
| 31/03/2020 | 35 | 1162,98699 | 22/04/2020 | 57 | 124510,788 | 14/05/2020 | 79 | 13330274,9 |
| 01/04/2020 | 36 | 1438,23834 | 23/04/2020 | 58 | 153979,529 | 15/05/2020 | 80 | 16485233,8 |
| 02/04/2020 | 37 | 1778,63514 | 24/04/2020 | 59 | 190422,82 | 16/05/2020 | 81 | 20386896,4 |
| 03/04/2020 | 38 | 2199,59576 | 25/04/2020 | 60 | 235491,371 | 17/05/2020 | 82 | 25211989,8 |
| 04/04/2020 | 39 | 2720,18774 | 26/04/2020 | 61 | 291226,576 | 18/05/2020 | 83 | 31179067,9 |
| 05/04/2020 | 40 | 3363,99146 | 27/04/2020 | 62 | 360152,977 | 19/05/2020 | 84 | 38558411,5 |
| 06/04/2020 | 41 | 4160,16821 | 28/04/2020 | 63 | 445392,618 | 20/05/2020 | 85 | 47684270,1 |
| 07/04/2020 | 42 | 5144,78104 | 29/04/2020 | 64 | 550806,454 | 21/05/2020 | 86 | 58970002,3 |
| 08/04/2020 | 43 | 6362,42831 | 30/04/2020 | 65 | 681169,237 | 22/05/2020 | 87 | 72926798,8 |
| 09/04/2020 | 44 | 7868,26372 | 01/05/2020 | 66 | 842385,789 | 23/05/2020 | 88 | 90186837,1 |
| 10/04/2020 | 45 | 9730,49454 | 02/05/2020 | 67 | 1041758,46 | 24/05/2020 | 89 | 111531916 |
| 11/04/2020 | 46 | 12033,4711 | 03/05/2020 | 68 | 1288317,91 | 25/05/2020 | 90 | 137928867 |

**Fig. 1.**
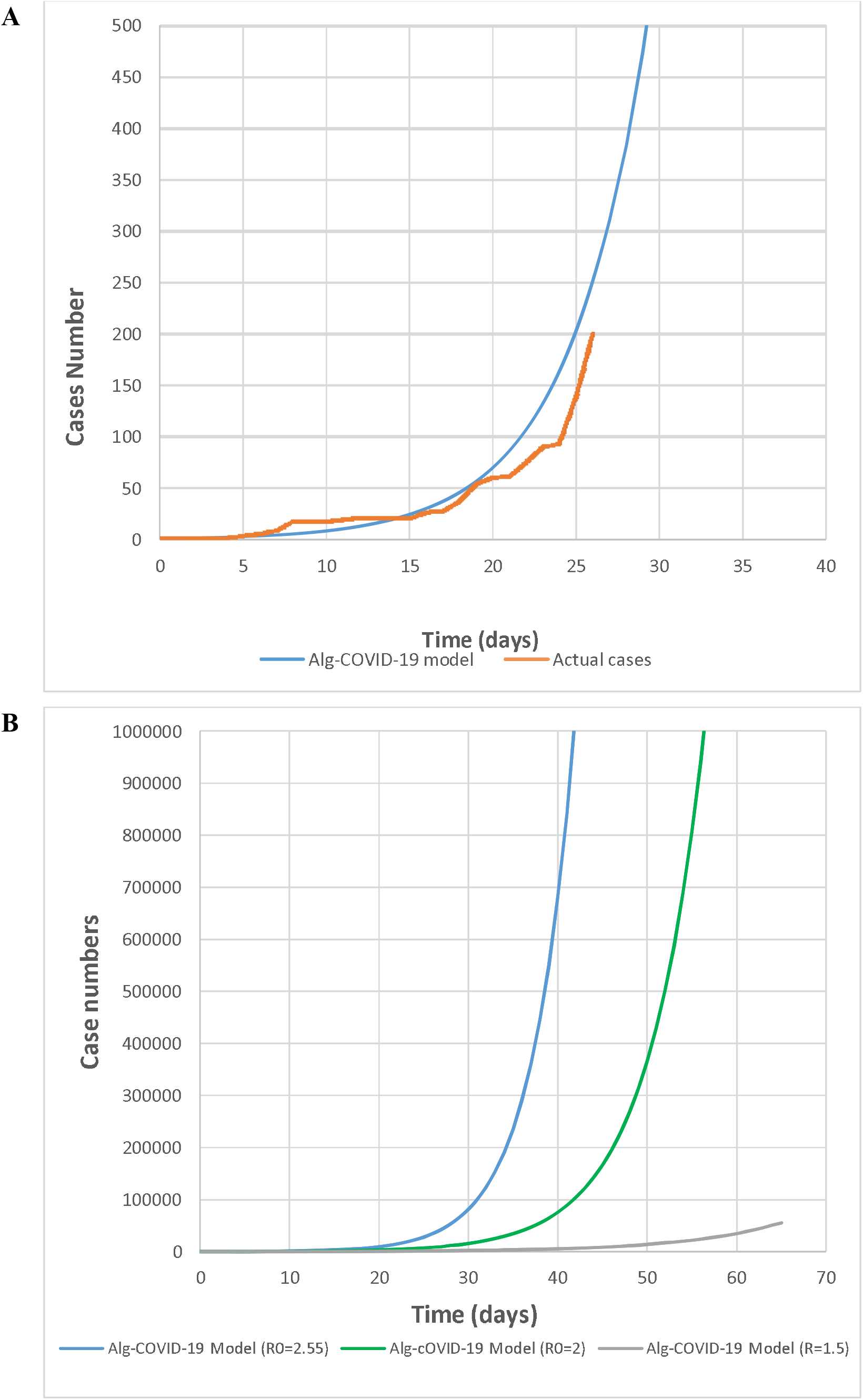
A: Alg-COVID-19 Model match to actual data (Cases number) of the first 26 days of epidemic. B: Alg-COVID-19 Model before and before (R0=2.55) and after mitigation (R0<2.55).

### Alg-COVIS-19 Model results

The estimations presented by this Model are based on the first 26 days data and cover the next months of COVID-19 epidemic in Algeria, according to that, the number of cases will exceed 1000 case on the 35^th^ day of the epidemic (March 31, 2020), 5000 on the 42^th^ day (April 7^th^) and it will double to 10000 on 46th day of the epidemic (April 11^th^), thus, exponential phase will increases continuously until reaching à herd immunity of 61% or a serious preventive measures are considered (Table 1; Fig.1.B).

## Discussion

Regarding the used parameters (R0 and SI) by Alg-CIVID-19 Model for the predictions, the estimated R0=2.55 (95% CI 2.15–2.93) until March 19, 2020, is very close to R0 that was found on the Chinese epidemic data R0=2,35 (95% CI 1·15–4·77) until February 4^th^, 2020, (Kucharski, et al. 2020), also, it is in the range of basic reproduction numbers of other studies mentioned below (Shen, et al. 2020, Read, et al. 2020, Chong, et al. 2020, Natsuko, et al. 2020, Tang, et al. 2020), on the other hand, the parameter that we cannot update and can affect the predictions is the serial interval (SI), we found two studies each one has different value that range from 4.4±3,17 (Chong, et al. 2020) to 7.5 days (95% CI, 5.3 to 19) (Li, et al. 2020), we used (SI=4.4) because of its standard deviation is not very large.

This model is valid only when the majority of the population is vulnerable to COVID-19 infection, since it does not take into account the herd immunity acquired ongoing epidemic. In case of R0 decreases over the next weeks of the epidemic, Alg-COVID-19 Model can be revalidated by recalculating a new effective reproductive number R1 (equation 3). However, this will happen only if the herd immunity rises spontaneously during the epidemic or the state act on one of the components of R0 by preventive measures (equation 2).

In the case of the Algerian COVID-19 epidemic, this Model can be valid for the first few months if the herd immunity remain low and susceptible population size is not changed, thus, it is applicable whenever a new regional outbreak begins with a naive population. On the other hand, if the state implement a good preventive policy according to the WHO guidelines (WHO.b 2020) and set up measures to reduce the effective contact rate, this will obviously reduce R0, so that, (R1) the effective reproduction number with control and preventive measures can be calculated to estimate the implemented preventive actions effectiveness, and Alg-COVID-19 Model can be updated with R1 values adapted to the new situation and its forecasts would correspond as closely as possible to the real epidemic growth (Fig.1.B). Also, it is obvious that the measures must be strong enough to reduce the basic reproduction number below 1 (R0 <1) to control the epidemic, otherwise, the epidemic will spread over time, which can also be a good strategy for maintaining the healthcare demand at a manageable pace.

According to Alg-COVID-19 Model results on Algeria data, the exponential phase 3 will probably announced after the 35^th^ day of the epidemic (1000 case) (March 31, 2020), consequently, the response to the epidemic will change from trying to curb the spread of the COVID-19 on the territory to a strategy that lead to mitigate the effects of the epidemic wave, so that, after R0 and SI update (Fig. 1.B), the Model can be used to predict the hospitalization and ICU or on a under ventilator patients numbers, and also, predict the number of deaths that remain the highest in the wold (Algeria: 8.5% until March 22^nd^) since the start of the pandemic, without any concrete explanation.

## Data Availability

All data refferd to in the manuscript is available

